# Advancing Understanding of Inequities in Rare Disease Genomics

**DOI:** 10.1101/2023.03.28.23286936

**Authors:** Jillian G Serrano, Melanie O’Leary, Grace VanNoy, Ingrid A Holm, Yarden S Fraiman, Heidi L Rehm, Anne O’Donnell-Luria, Monica H Wojcik

## Abstract

**Purpose:** Advances in genomic research have led to the diagnosis of rare, early-onset diseases for thousands of individuals. Unfortunately, the benefits of advanced genetic diagnostic technology are not distributed equitably among the population, as has been seen in many other healthcare contexts. Even quantifying and describing inequities in genetic diagnostic yield is challenging due to variation in referrals to clinical genetics practices and other barriers to clinical genetic testing.

**Methods:** The Rare Genomes Project (RGP) at the Broad Institute of MIT and Harvard offers research genome sequencing to individuals with rare disease who remain genetically undiagnosed through direct interaction with the individual or family. This presents an opportunity for diagnosis beyond the clinical context, thus eliminating many barriers to access.

**Findings:** An initial goal of RGP was to equalize access to genomic sequencing by decoupling testing access from proximity to a major medical center and physician referral. However, our study participants are overwhelmingly non-disadvantaged, as evidenced by their access to specialist care and genetic testing prior to RGP enrollment, and are also predominantly white.

**Implications:** We therefore describe our novel initiative to diversify RGP enrollment in order to advance equity in rare disease genetic diagnosis and research. In addition to the moral imperative of medical equity, this is also critical in order to fully understand the genomic underpinnings of rare disease. We utilize a mixed methods approach to understand the priorities and values of underrepresented communities, existing disparities, and the obstacles to addressing them: all of which is necessary to promote equity in future genomic medicine initiatives.

## Introduction

Remarkable advances in genomic sequencing have transformed rare disease diagnosis (1, 2).. The ability to search for pathogenic variants across the genome have led to the identification of a molecular diagnosis for many individuals and the discovery of thousands of disease genes (1, 2). This has a profound and multifaceted impact on these individuals and their families, in addition to broader implications for the larger rare disease community towards understanding the genomic landscape of these conditions. At the level of the patient, the psychosocial challenges of remaining undiagnosed may be alleviated by a molecular diagnosis, allowing for increased understanding, feelings of empowerment and control, and connection with other people with the same condition for further support (14, 15). Unfortunately, the benefits of these advances in genomic technology are not distributed equitably, as has been seen in many other aspects of healthcare (3, 4). Rates of genetic diagnosis are lower in minoritized individuals (16, 17).

Additionally, multiple prior studies of genomic sequencing, its diagnostic yield, and its clinical and psychosocial impact involve primarily white participants with a relatively high socioeconomic status (4, 18, 19). There are many structural and systemic barriers to a genetic diagnosis among diverse populations (5, 6), including failure to suspect or recognize genetic disorders in non-white individuals (7), decreased referral rates for clinical genetic testing, insurance challenges, difficulty interpreting test results given lack of ancestry diversity in the reference genome (8), and low referral rates to rare disease research programs when standard clinical evaluation is unsuccessful (39).

These inequities in access to genomic sequencing are not the result of lower incidence of genetic disorders in non-white populations, but highlight systemic and structural barriers in access to appropriate health services. Indeed, quantifying and defining inequities in genetic diagnostic yield has also been challenging, particularly because a genetic diagnostic evaluation is never even initiated for many individuals with limited access to healthcare, as we have previously demonstrated (35). This may result in fewer diagnoses identified in minoritized populations, a longer time to diagnosis (40) and a lack of diversity in genomic research studies (4, 9). In addition, the features leading to effective implementation of genomic medicine for underserved communities remain poorly understood (22). This, in turn, impedes optimal development of genomic medicine programs and perpetuates inequities.

The Rare Genomes Project (RGP) is an ongoing study offering research genome sequencing (GS) that is free to participants for whom the clinical route to testing has been unsuccessful. A goal of RGP is to equalize access to genomic sequencing by decoupling testing access from proximity to a major medical center and physician referral and by removing cost and insurance coverage as barriers to access. However, our study participants who have self-reported race and other sociodemographic features remain overwhelmingly white (>90%), well-educated (>90% of adults with some college attendance or higher) and well-resourced, as evident from their testing history prior to RGP enrollment and self-reported household income (>60% reporting annual income >$90,000/year and only 5% below the federal poverty line). Although insurance is not a barrier in research, the process required to enroll in a research study, including RGP, and have genomic sequencing performed is still fraught with multiple barriers to access. Diverse and population-representative genomic studies that include individuals from multiple ancestries will ideally augment the ability to identify molecular diagnosis for individuals with rare disease. as has been seen already in research on complex traits (10). As such, research GS with a specific focus on broad and diverse enrollment presents a unique opportunity to not only improve genetic diagnoses for under-represented individuals. by potentially eliminating access barriers, but also to generate data to improve diagnostic likelihood for all. We therefore plan to study the impact of approaches to address the systemic and structural barriers to genetic testing and hypothesize that doing so will lead to an increased number of diagnoses in underserved communities. Coupling qualitative approaches also presents an opportunity to understand the priorities and values of historically medically-underserved communities and the existing inequities perpetuated by structural barriers.

## Methods

### Overview of Study Design

This study was developed with input from key stakeholders across multiple domains, including clinicians, genomic researchers, genetic counselors, and individuals with rare disease or their family members, including partnerships with many patient advocacy organizations. We have three primary aims: 1) to diversify recruitment, 2) understand the process and context of implementing genomic medicine for rare disease diagnosis, and 3) investigate the value of a diagnosis for underserved populations. Inclusion criteria include the presence of a condition that is likely to be monogenic, the ability to provide samples for genetic testing, and residence in the United States. Exclusion criteria include a confirmed genetic diagnosis or a condition unlikely to be monogenic. In addition to genomic data (first aim), we will collect and analyze data regarding the implementation of our process to diversify RGP enrollment (second aim) and survey data regarding diagnostic impact for participants (third aim).

### The Rare Genomes Project

Overview: The typical process of RGP enrollment begins with the affected individual or their parent/guardian self-referring using an application found on our website (Figure 1), often connected to us via social media or at the recommendation of another individual, patient advocacy organization, or provider who knows about the project. Participants can access additional information by reviewing information on the website, including a brief participation overview video, and by visiting the RGP Facebook page that is maintained by study staff. An internal application review process is then performed by two clinicians to determine the eligibility of these applicants for enrollment. For cases that are accepted, RGP study staff contact the affected individual or parent/guardian to explain the study and obtain informed consent for enrollment in the RGP study protocol from affected individuals and their family members. Finally, blood samples from the affected individuals and biologic parents (when available) are obtained for GS and analysis. If a likely diagnosis is found, the causal variant(s) are confirmed in a CLIA-certified lab (cost covered by the study) and returned through the participant’s local physician.

**Figure 1.** The Rare Genomes Project homepage. From this website, participants may click a link to self-apply or apply on behalf of their child for consideration of study enrollment.

Consent: If applicants are eligible to participate, they are contacted via email and prompted to schedule a consent appointment through their online dashboard within the RGP website. If the participant does not have access to online resources including their dashboard, an RGP staff member contacts the participant via phone to schedule a consent appointment. The consent process is typically 30-45 minutes in length and held over Zoom. If the participant does not have access to Zoom, the consent discussion occurs by phone.

Data and sample collection: Once enrolled, a blood draw collection kit is sent to the participant’s mailing address. The participant or RGP staff identifies a phlebotomy site and has a blood sample drawn and returned for GS. Medical records are collected after enrollment to be used at time of analysis of the data. Participants or parents/guardians provide RGP staff with the medical records directly, or sign release forms for study staff to obtain records from relevant facilities.

Sequencing and analysis: Following enrollment and sample procurement, GS is performed by the Genomics Platform (GP) at the Broad Institute and analyzed by the RGP analysis team through the Broad Institute Center for Mendelian Genomics.

Reporting and disclosure: Diagnostic variants and candidates of high clinical suspicion that are approved by RGP leadership (which includes board-certified clinical geneticists and molecular diagnosticians) undergo CLIA-certified confirmation and are returned to the families in coordination with their local provider. If families do not have a local provider willing or comfortable ordering the confirmation test, a genetic counseling telemedicine visit is offered. Variant classifications are shared in ClinVar to increase public knowledge about pathogenic variants in understudied populations. De-identified genomic data from all cases is also shared with other researchers through the National Human Genome Research Institute’s AnVIL.

### Population and recruitment

The targeted enrollment for this prospective cohort study is 50 participants who are currently underrepresented in the RGP cohort. The following criteria will be used to determine underrepresentation: non-white race, Hispanic or Latinx ethnicity, limited English proficiency, household income under the federal poverty line, parental education high school level or less, and primary residence in a non-metropolitan area (Table 1). These criteria are identified in potential participants in multiple ways: RGP applicants are asked to self-identify their race and ethnicity at the time of initial application, or referring clinicians may identify the race and ethnicity for the individuals whom they refer; enrolled participants also provide additional demographic data via our baseline RGP survey (which participants complete online, on paper, or via interview).

**Table 1.** Underrepresentation in the Rare Genomes Project cohort.

| **Category** | **Proportion prior to the current study** |
| --- | --- |
| Non-white race | 8.0% |
| Hispanic or Latinx ethnicity | 8.5% |
| Limited English proficiency | 0.0%<sup>a</sup> |
| Household income under the federal poverty line | 5.0% |
| Parental education high school level or less | 3.8% |
| Primary residence in a non-metropolitan area | 15% |

<sup>a</sup>English proficiency was one of the original inclusion criteria for this study

Recruitment strategies will include outreach to clinicians who may care for a large population of underrepresented individuals, in addition to community partnerships. A short physician referral form that typically takes <5 minutes to complete has been created for physicians to refer patients who would typically face barriers through the online RGP application process. Additional outreach efforts include establishing relationships with rare disease organizations that focus on increasing access to genomic medicine for minoritized communities.

### Implementation of enhanced recruitment and enrollment support

The virtual nature of RGP makes participating in the study feasible for those who have access to online resources such as a computer or smartphone, along with internet or cellular data access. However, since flexibility within the RGP study design is needed for underserved populations, we identified multiple barriers to enrollment, including limited English proficiency, difficulty accessing online resources, lack of transportation, and inability to communicate with RGP study staff during normal working hours. The following modifications will be implemented into the enrollment process to address barriers within RGP (Figure 2): (1) Outreach to providers to identify individuals with rare diseases who are unable to access clinical genetic testing. (2) Application assistance by RGP staff for any participants who have difficulty accessing online resources, rather than relying on self-referral via our website. The online application will be completed over the phone with the participant and a Spanish-speaking RGP coordinator or an interpreter for other languages if needed. Alongside this, RGP staff will make themselves available outside normal working hours to accommodate participants who need assistance during evenings and weekends. (3) Increased access to interpreter services and translated study materials. While the RGP study initially launched in English, the protocol was amended to eliminate the English language requirement and the website and recruitment materials have been fully translated into Spanish, with other languages to follow. (4) Mobile phlebotomy services for participants unable to travel to a clinical lab to provide a sample for sequencing (many local labs close by 1pm, presenting a potential barrier to access) (5) Finally, as certain families may have ambivalence about genetic testing in general, we will continue to discuss questions and concerns with prospective participants, which will also inform our process. We anticipate that these strategies will allow us to increase access to RGP by addressing some of the structural barriers to race, language proficiency, geographic location, and socioeconomic status.

**Figure 2.**
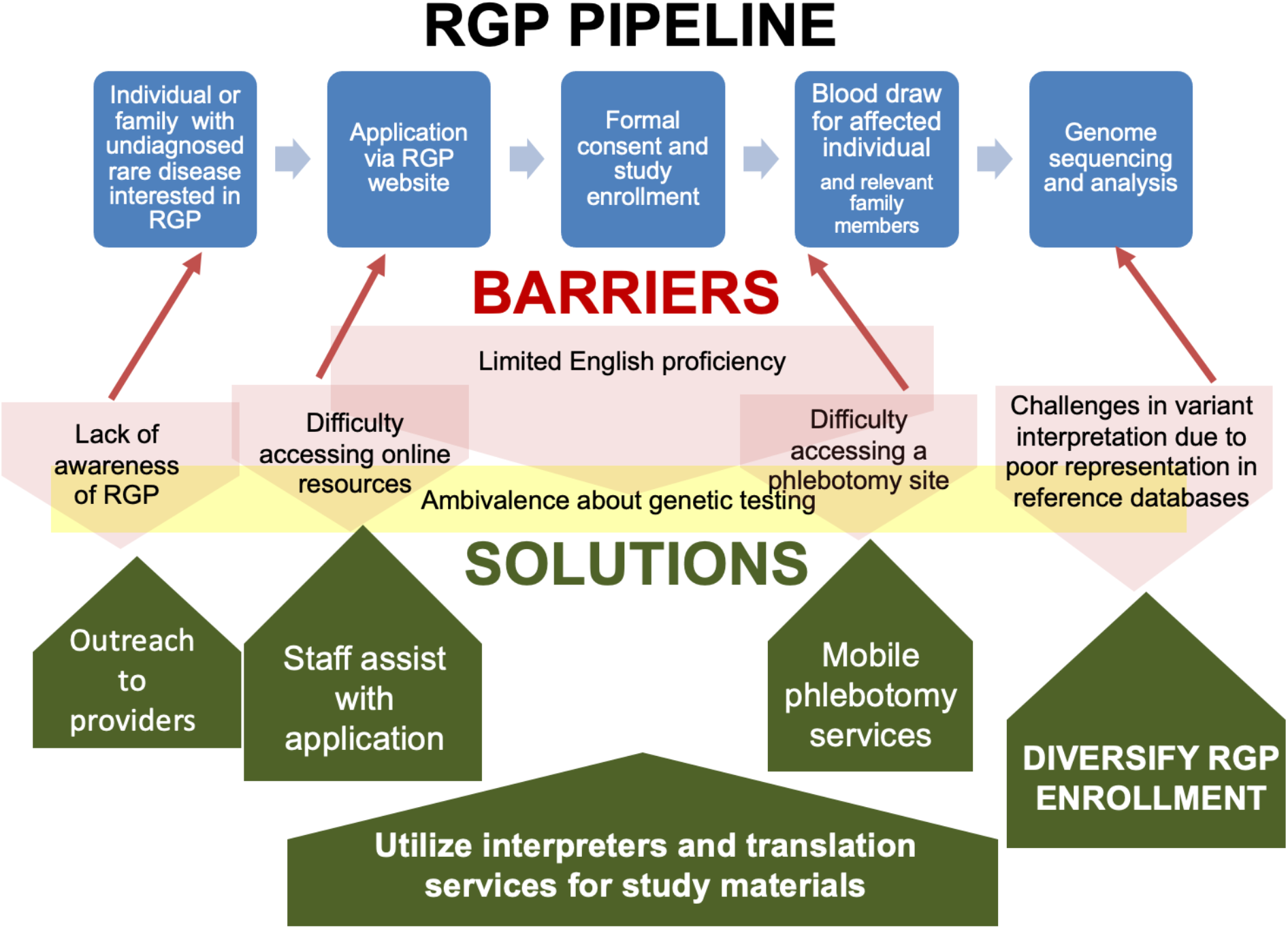
Rare Genomics Project enrollment process, barriers, and proposed solutions. The original process is represented by the blue squares, barriers by the red and yellow shaded shapes, and solutions by the green arrows.

### Evaluation of the intervention

To evaluate the process and context of implementing genomic medicine for rare disease diagnosis in diverse populations, we will employ a mixed-methods approach informed by Consolidated Framework for Implementation Research (CFIR) constructs as portrayed in a recent taxonomy (27), used to design our outcome measures in order to generate valid empirical data. The implementation outcomes of adoption (measured at the level of the provider) and acceptability (measured at the level of the consumer, in this case, the study participant) are measured in this project, where adoption refers to the uptake of the intervention and acceptability refers to how agreeable the intervention is to the participant (27). The referent for these outcomes will be the RGP process from enrollment to sequencing, analysis, and return of results (Figure 3). We will evaluate participant features associated with lack of completion of the RGP process related to our implementation outcomes.

**Figure 3.**
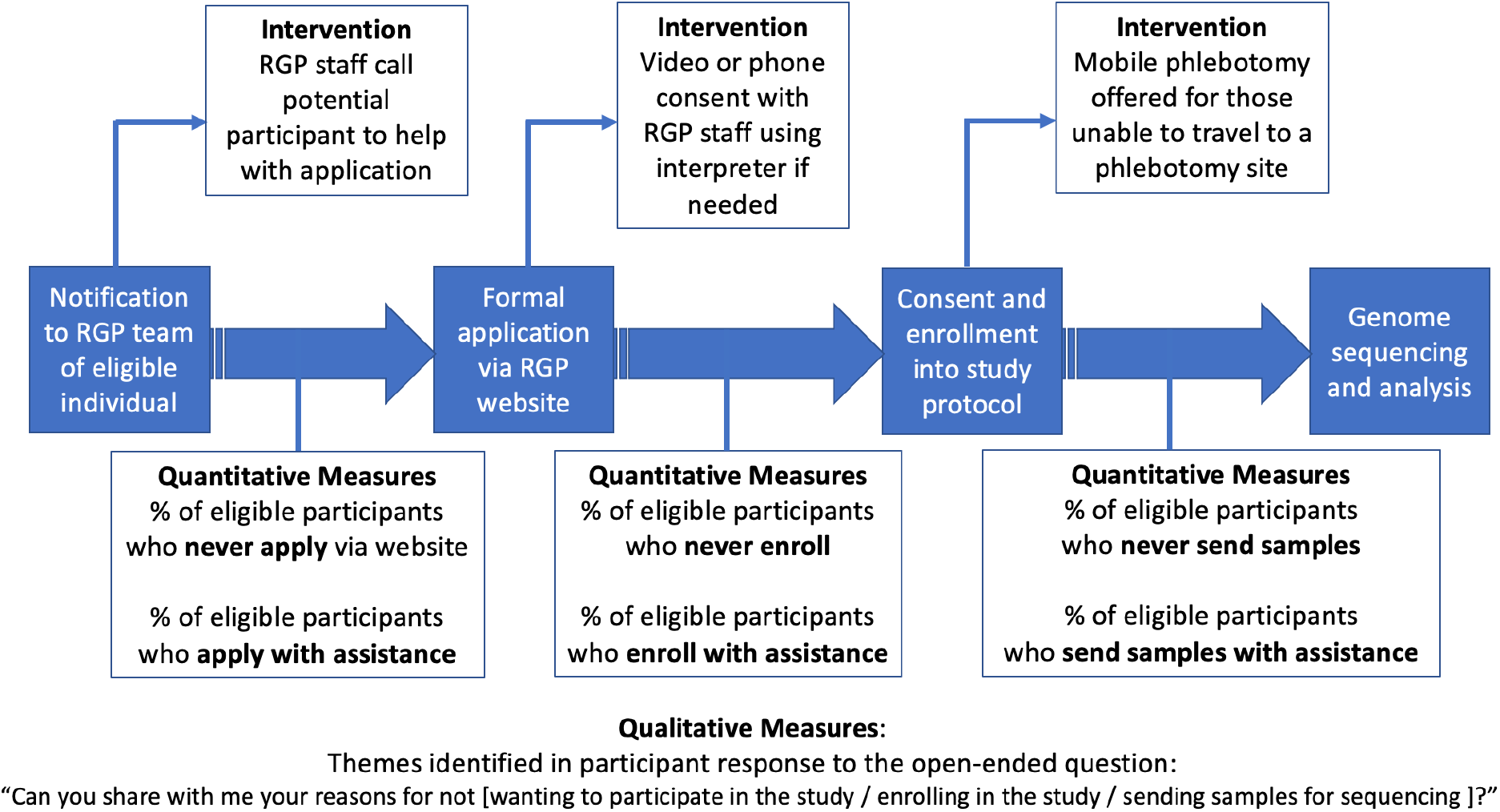
The Rare Genomes Project process, interventions to improve diversity, and implementation outcomes measures. Interventions are presented in text boxes above the current workflow (in blue), and outcomes measures are presented in text boxes below.

Quantitative measures include proportions of individuals who are able to progress through the various stages of the study. For the qualitative component of the study, RGP study staff will contact prospective participants who have been referred but do not apply via our website, who apply but do not complete enrollment, or who have enrolled but do not provide samples for sequencing (see below). We will use this information to address unforeseen barriers that are identified.

Referral/application stage: We will note the number of individuals who complete this process with assistance or who do not proceed further. Those who elect not to proceed further will be offered the opportunity to share their experience in an open-ended manner. Enrollment stage: We will note the number of prospective participants who decline enrollment and those who require the assistance of interpreter services to enroll. Those who decline enrollment will be asked to share their reasoning as above.

Sample collection stage: If a sample has not been received three months after enrollment, we will contact the participant or family to determine why samples have not been sent and whether mobile phlebotomy would be useful. If the participant would like mobile phlebotomy services, we will arrange for this and re-contact participants again if samples have not been received three months after mobile phlebotomy is offered. We will note the number of participants who require mobile phlebotomy to provide samples and those who never provide samples. Those who decline to provide samples at any time will be asked to share their reasoning as above.

In this way, adoption will be assessed by determining the proportion of referrals/applicants that do not enroll in the study or enroll but do not submit samples for sequencing within the first six months after enrollment. Acceptability will be assessed by evaluating the reasons that potential participants do not proceed from referral to receipt of samples for GS and the motivations for pursuing a diagnosis.

### Understanding priorities of study participants

To evaluate the priorities, values, and impact of a diagnosis for underrepresented individuals and families, we developed baseline and post-diagnosis follow-up survey tools that address multiple domains of diagnostic impact and are informed by CFIR constructs (Table 2). This survey has been in use within RGP for the past year and has been well-received. Surveys will be administered to enrolled participants or their parents/guardians. The study surveys have been translated into Spanish for the purposes of this project. For additional languages, we will offer verbal administration of the surveys using interpreter services or translation of the written questions into additional languages for study participants who speak languages other than English or Spanish. RGP participants or their parents/guardians will also be asked to complete the Perceived Stress Scale (PSS), a validated instrument that is also available in multiple languages (28). The PSS is a 10-item survey that asks participants to respond on a 5-point Likert scale from “never” to “very often” regarding certain thoughts and feelings over the past month. A total score is calculated, with lower scores reflecting less stress, and comparisons may be made to population averages.

**Table 2.** Domains included in study surveys are baseline and follow-up

| **Variable Domains** | **Baseline** | **Follow-up** | **CFIR Construct** |
| --- | --- | --- | --- |
| **Participant Characteristics** |  |  | **Personal attributes** |
| Socio-demographics* | X |  |  |
| **Utility of Genetic Diagnosis** |  |  | Knowledge and Beliefs about the Intervention |
| Overall importance | X | X |  |
| Understanding | X | X |  |
| Responsibility | X | X |  |
| Worry | X | X |  |
| Preparation for the future | X | X |  |
| Better treatment | X | X |  |
| Social support | X | X |  |
| Indirect healthcare costs | X | X |  |
| Decision to have more children | X | X |  |
| Planning for next pregnancy | X | X |  |
| **Perceived Stress Scale** | X | X | Personal Attributes |

\* Including race, ethnicity, household income, education level

The RGP baseline survey and PSS will be offered upon enrollment and the follow-up survey and PSS repeated at 3- and 12-months post diagnosis (if one is found via GS) to evaluate changes in priorities, values and stress level. Surveys may be completed online via REDCap (29) (preferred) or on paper via mail. Modifications for this study will include administering the survey via an interview for those who are unable to submit it online or via mail. Our preliminary data has demonstrated the feasibility of using this survey tool, with 62% of enrolled participants completing the baseline survey, and 89% of eligible participants completing the 3- and 12-month follow-up surveys. While race, ethnicity, and zip code are collected in the RGP application, the remainder of the sociodemographic criteria used to determine underrepresentation will be collected via the surveys.

### Analysis

Genomic analysis: We will analyze the GS data to identify molecular diagnoses under the assumptions that the affected individual has a severe, rare, Mendelian condition, and we clinically-confirm our top candidates identified from genome sequencing. Our interpretation of gene-disease association is guided by the Clinical Genomic Resource (ClinGen) framework (30) and variants are classified according to criteria defined by the American College of Medical Genetics and Genomics (ACMG) (31). We return pathogenic, likely pathogenic, and variants of uncertain significance (VUS) of high interest in established disease genes, along with VUSs of high interest in candidate disease genes. We consider a case “solved” if a variant is clinically validated (covered by study funds, typically CLIA Sanger confirmation) and the local physician agrees with the interpretation after evaluating the genetic test results and the patient.

Diagnostic yield: We will compare the diagnostic yield in our cohort of 50 RGP participants who qualify as underserved or historically underrepresented (based upon our six criteria outlined in Table 1) to the yield in the larger RGP cohort. Additional analyses will include a) the proportion of diagnoses requiring GS (versus those that could have been found using other common clinical tests such as chromosomal microarray, karyotype, or targeted gene panel), b) the proportion of diagnoses involving previously-known versus novel disease genes and previously-known vs novel variants, and c) the phenotypic characteristics of each population. Phenotypes are characterized using the Human Phenotype Ontology (HPO) (32), and we will identify differences in number and types of HPO terms associated with the underserved and non-underserved cohorts.

Analysis of intervention success: We will compare the overall proportions of RGP participants across the entire study period who fall into our six measured socio-demographic categories both before and after the implementation of the described measures. This will indicate whether or not we have achieved our goal of increasing RGP diversity. Subsequent analyses will focus on the process and context of our implementation measures, evaluating the implementation outcomes of adoption and acceptability of GS for a diverse cohort of individuals with rare disease (Table 3). The first implementation measure will quantify the percentages of individuals eligible for RGP who do not complete (a) the application, (b) enrollment, or (c) return of samples for GS. These outcomes will be further explored using a mixed-methods approach involving quantitative data from our RGP study surveys and qualitative data obtained from the open-ended question posed to participants who do not proceed from eligibility to application, from application to enrollment, or from enrollment to provision of samples for GS.

**Table 3.** Implementation Outcomes

|  | Quantitative Measures | Qualitative Measures |
| --- | --- | --- |
| Adoption | Proportion of referred participants who fully enroll<br>Proportion of enrolled participants who are sequenced<br>Proportion of participants belonging to at least one underserved category |  |
| Acceptability | Differences in motivations for pursuing GS and in perceived impact of genetic diagnosis | Reasons not to participate and free-text responses on RGP survey |

Analysis of psychosocial impact of GS: Survey data will be collected using REDCap (29) and downloaded for descriptive statistical analyses. The primary outcome will be the perceived overall importance of a genetic diagnosis, measured on a 5-point Likert scale (“not at all important” to “extremely important”). Secondary outcomes include other domains of diagnostic utility, also measured on a 5-point Likert scale. An additional secondary outcome is the mean perceived stress score determined by the 10-item PSS (28). We will determine the effects of participant characteristics on this primary outcome by performing a multiple linear regression analysis with perceived importance of a genetic diagnosis as the dependent variable and socio-demographic features as covariates. We will test variables individually and construct regression models to evaluate the interaction between these and other covariates such as clinical features of the affected child. For our secondary outcomes, we will similarly evaluate the influence of socio-demographic features on the various domains of perceived diagnostic utility and on the perceived stress score. Additionally, we will evaluate the impact of a diagnosis by comparing the domains of diagnostic utility and perceived stress at baseline to these same features both immediately (3 months) and longer-term (12 months) post-diagnosis for those who receive a molecular genetic diagnosis.

## Anticipated Results

The first goal of this study is to increase genetic diagnoses for medically-underserved populations through outreach and enhanced enrollment support. Through expansion to non-English speakers, a personalized, supportive approach to participation, and targeted outreach to populations currently underrepresented in RGP, we hope to provide access to GS to a more diverse cohort. We will measure the diversity of the RGP cohort related to several demographic categories that are currently underrepresented both in our cohort and in previous publications (25, 26) (Table 1). The diagnostic yield from GS will be compared between this cohort (N = 50) and RGP participants who do not meet one of the 6 criteria listed above and are not considered to be historically underserved or underrepresented. Our framework conceptualizes underrepresentation as a result of structural and systemic discrimination. As such, we hypothesize that there is no molecular or genetic basis of molecular underdiagnosis in underrepresented populations. In fact, we hypothesize that there will increased diagnostic yield within our diverse and underrepresented cohort compared to those that do not meet one of the 6 criteria of underrepresentation as defined by the study because of referral bias, as we have previously shown (35). A clear benchmark for success for this study will be an increase in the racial, ethnic, and socioeconomic diversity of the RGP study cohort.

A second goal is to evaluate the process and context of implementing genomic medicine for rare disease diagnosis in diverse populations. A crucial outcome from this study will be an analysis of the process involved in successful rare disease research enrollment, sample acquisition, analysis, and return of results for underserved populations. Outcomes of adoption will be measured based on the percentage of referred participants who fully enroll, percentage of enrolled participants who are sequenced, and the percentage of participants in ≥1 underserved category. Outcomes of acceptability will be measured by evaluating the reasons participants do not proceed from referral to GS.

Our final goal is to evaluate the priorities, values and impact of a diagnosis for underrepresented individuals and families. We will describe the motivations for seeking a diagnosis and diagnostic impact (both psychosocial and related to healthcare utilization) in this cohort of 50 individuals with comparison to our overall RGP study population. We hypothesize that the current overrepresentation of white, well-resourced participants is the result of access barriers rather than differences in the perceived benefit of a genetic diagnosis, though prior research suggests that perceived utility of genetic testing may vary by cultural context (37).

## Discussion

The Rare Genomes Project is an innovative study both related to the ability for prospective participants to self-refer and participate from anywhere in the United States without receiving care at a major academic medical center. In addition, diagnostic GS results are returned, as well as promising candidate disease genes, directly to RGP participants in coordination with their local medical care team (in addition to our genetic counseling team). The goal of RGP is to empower families with genomic understanding of their own rare conditions. However, due to lack of diversity in genomic research studies for individuals with rare disease, the optimal approach for implementation of genomic medicine, particularly for medically-underserved and minoritized populations, remains unknown and under-realized (38). We therefore present our approach to devote resources to facilitate access for these populations to our ongoing research study, while concurrently analyzing not only the diagnostic, clinical, and psychosocial impact of GS but also the approach itself, via our implementation outcomes.

Like many research studies, RGP was originally designed to be accessible by any person. However, structural barriers including the need to navigate online resources and speak English contributed to the inequitable access to RGP and ultimately led to the lack of diversity within the study, as prior research has demonstrated that language barriers as well as dependence upon referrals from subspecialists, such as clinical geneticists, obstructs access to rare disease research (39). To increase the outreach and engagement with underserved families, we are modifying the typical RGP process to expand the use of clinician referral forms, provide assistance over the phone rather than relying on the online application and Zoom for consent discussions, utilize mobile phlebotomists for those with transportation barriers, expand our availability to accommodate those with long working hours, overcome language barriers through amendment to study eligibility and utilization of interpreter services, and discuss questions and concerns with prospective participants. Through this implementation process, we hope to improve the participant experience and make our study accessible to those who would not have otherwise participated in RGP. With outreach to providers, particularly those who treat a large underrepresented population, we aim to spread awareness about our study. One potential problem that we foresee is suboptimal enrollment of historically underserved populations to our study. Issues of trust regarding enrollment in research, particularly genomic research, have been identified related to minoritized racial groups both in the United States and elsewhere (33, 34). As enrollment in RGP provides a service to participants in performing GS and identifying diagnoses that have been difficult for them to access previously, we hope that our approach may begin the process of reparative justice to a therapeutic relationship between research and participant that has been marred by past injustice, disenfranchisement, and medical racism. As RGP staff works closely with the individuals and families to help overcome challenges, we hope to build trust with the participants and improve the experience of the diagnostic odyssey.

We anticipate that the diagnostic yield of GS in underrepresented populations may be higher than the current cohort, as our prior research into diagnostic yield in underserved populations has identified a higher odds of diagnosis in minoritized populations - which we attributed to referral bias for more severe phenotypes (35). Furthermore, if our strategies to diversify enrollment are successful, this would provide needed insight on successful approaches to reach underserved populations and increase access to genomic medicine for other rare disease researchers and potentially for those working in a clinical setting as well. The design of this study is such that even if participation is not increased significantly, we will generate valuable insight into any shortcomings of our implementation process.

Prior research suggests that implementation science has potential to address the barriers and challenges around health inequities in genomic medicine (36). However, prior initiatives related to genomic medicine implementation have focused on individual rather than contextual factors and less than 2% invoked an implementation science framework, without which it is difficult for results to be generalizable and informative; additionally, individual-level interventions to address inequity are often unsuccessful (22). Based on a literature review using the Centers for Disease Control and Prevention’s Public House Genomics Knowledge Base to examine implementation science in genomic medicine, the racial and ethnic composition of study populations are often underreported, limiting the ability to contextualize and generalize study outcomes (22). Additionally, prior research related to inequities in genomic medicine have focused primarily on cancer or prenatal genetics and have not been centered around rare disease diagnosis in children and adults (24). Therefore, this presented protocol seeks to address an understudied area of rare disease genomic research.

## Conclusions

We present our approach to address inequities in access to genomic research for individuals with rare disease. Our plan to measure not only diagnostic yield for diverse populations but also to generate empiric data regarding implementation outcomes is crucial to maximize the power of the conclusions from this project as well as to inform further efforts in the field. Our focus on implementation outcomes rather than solely on clinical outcomes provides valuable insight into the context and process of achieving the desired clinical outcomes and will allow for adjustment of approach if the desired clinical outcomes are not achieved. Even a highly effective clinical intervention such as GS may not be successful in “real world” practice if various implementation factors, such as the acceptability and adoption that we are measuring, are not optimized. As precision medicine continues to rapidly develop, ensuring equitable access to these technological advances is paramount.

## Data Availability

All data produced in the present work are contained in the manuscript.

## Acknowledgements

The authors would like to express their gratitude to all the families who participate in our ongoing research.

